# Scabies epidemiology in health care centers for refugees and asylum seekers in Greece

**DOI:** 10.1101/2022.01.10.22268996

**Authors:** Christina Louka, Emmanouil Logothetis, Daniel Engelman, Eirini Samiotaki Logotheti, Spyros Pournaras, Ymkje Stienstra

## Abstract

**Background:** Scabies is a global health concern disproportionally affecting vulnerable population such as refugees and asylum seekers. Greece is a main geographical point of entry in Europe for refugees, but epidemiological data on scabies in this population is scarce. We aimed to evaluate the epidemiology of scabies, including trends over the study period.

**Methodology/Principal findings:** Data were collected from June, 2016 to July, 2020, using the surveillance system of the Greek National Public Health Organization. Staff at health centers for refugees/asylum seekers compiled daily reports on scabies and other infectious diseases. Observed proportional morbidity for scabies was calculated using consultations for scabies as a proportion of total consultations.

There were a total of 13118 scabies cases over the study period. Scabies was the third most frequently observed infectious disease in refugees/asylum seekers population after respiratory infections and gastroenteritis without blood in the stool. The scabies monthly observed proportional morbidity varied between 0.3% (August 2017) to 5.6% (January 2020). Several outbreaks were documented during the study period. An increasing number of cases was observed from October 2019 until the end of the study period, with a peak of 1663 cases in January 2020, related to an outbreak at one center. Spearman correlation test between the number of reported scabies cases and time confirmed an increasing trend (ρ=0.67).

**Conclusions/Significance:** Scabies is one of the most frequently reported infectious diseases by health care workers in refugee/asylum seekers centers in Greece. Consultations for scabies increased over time and there were several outbreaks. The current surveillance system effectively detects new cases in an early stage. Public health interventions, including mass drug administration, should be considered to reduce the burden of scabies in refugee/migrant populations.

**Author summary:** Scabies is a skin disease caused by ectoparasitic mite *Sarcoptes scabiei* var. *hominis*. A person infected by scabies manifests symptoms like itching and skin rash, while the most complicated forms of the infection can lead to bacterial infections and sepsis. Scabies is a global health concern and as of 2017, it is officially included in the portfolio of conditions prioritized by the Department of Neglected Tropical Diseases, according to the World Health Organization. Furthermore, scabies can disproportionally affect vulnerable population, such as refugees and asylum seekers. Greece is one of the main points of entry in Europe for refugees. However, the prevalence of scabies amongst refugees in Greece has not been thoroughly investigated. In this study, we utilized data weekly reported within the epidemiological surveillance in refugees/asylum seekers centers of the Greek National Public Health Organization, in order to obtain a clear overview of scabies epidemiology and identify any trends over time. Our findings confirmed the high prevalence of scabies among refugees and identified a clear trend of significant increase over time. Further public health interventions, like mass drug administration could help restrain the dissemination of the disease and lower its burden among refugees.

## Introduction

Scabies is a skin disease caused by infestation with ectoparasitic mite *Sarcoptes scabiei* var. *hominis*. The main clinical manifestations include mild to severe itching and a skin rash consisting of small papules, nodules and vesicles. Scabies can be further complicated, leading to secondary bacterial skin infections, invasive bacterial infections, and sepsis [1].

Scabies is a global health concern affecting an estimated 200 million people worldwide. It is endemic in many low- and middle-income countries, particularly those with hot, tropical climates, and it disproportionally affects children and vulnerable population [2,3]. Overcrowded conditions, low income and limited access to treatment are factors associated with increased prevalence. In 2017, the World Health Organization officially included scabies in the portfolio of conditions prioritized by the Department of Neglected Tropical Diseases.

Ongoing armed conflict, political turmoil and financial instability has led to an increased influx of asylum seekers and refugees into Europe [4]. Several studies have demonstrated the increased prevalence of scabies amongst asylum seeker populations in Europe [5-7]. Greece is a major geographical point of entry for refugees, mainly originating from Afghanistan, Syria, Iraq, and the African continent. According to the United Nations High Commissioner for Refugees, approximately 1.2 million refugees have entered Greece through sea or mainland, from beginning of 2015 until end of June of 2021 [8]. After their arrival, refugees are appointed in designated centers located on several Aegean islands and the Greek mainland.

Specialized medical personnel are appointed in Points of Care (PoC) within refugees/asylum seekers centers and are responsible for the healthcare, medical examination and subsequent treatment of the refugees/asylum seekers. The National Public Health Organization (NPHO) provides directions and instructions for the management of a range of communicable diseases, including scabies to medical personnel. For scabies, instructions cover the detection, treatment, individual and environmental preventive measures, and management of scabies outbreaks [9].

Scabies is not a mandatory reportable disease for general population in Greece. Nevertheless, surveillance amongst refugees/asylum seekers in PoC is conducted by the NPHO. Clinical scabies cases are documented throughout the country and reported on a weekly basis within the surveillance system for PoC. However, epidemiological data regarding scabies in refugees and asylum seekers in Greece is scarce. We aimed to evaluate the epidemiology of scabies in refugees/asylum seekers centers in Greece, to investigate changes over time and factors relating to these changes and to compare the epidemiology of scabies with other reported infectious diseases.

## Methods

### Identification and assessment of refugees/asylum seekers

Refugees/asylum seekers may enter Greece by sea or land. As soon as practicable after arrival, NPHO personnel conduct an initial, preliminary medical examination and identify those in need of immediate medical assistance and/or hospital admission. Medically stable refugees/asylum seekers complete registration and identification procedures and are relocated to one of thirty-nine Regional Units [10]. These include six ‘Reception and Identification Centers’, located across the sea and land borders, where newly arrived refugees are documented and identified. Thirty-one ‘Facilities of Temporary Reception’ where refugees are relocated whilst undertaking in the process of seeking asylum, are on the Greek mainland (Figure 1). When the criteria to apply for asylum are not met, people may remain within refugees/asylum seekers centers for a prolonged period of time, until their situation is reevaluated.

**Figure 1.**
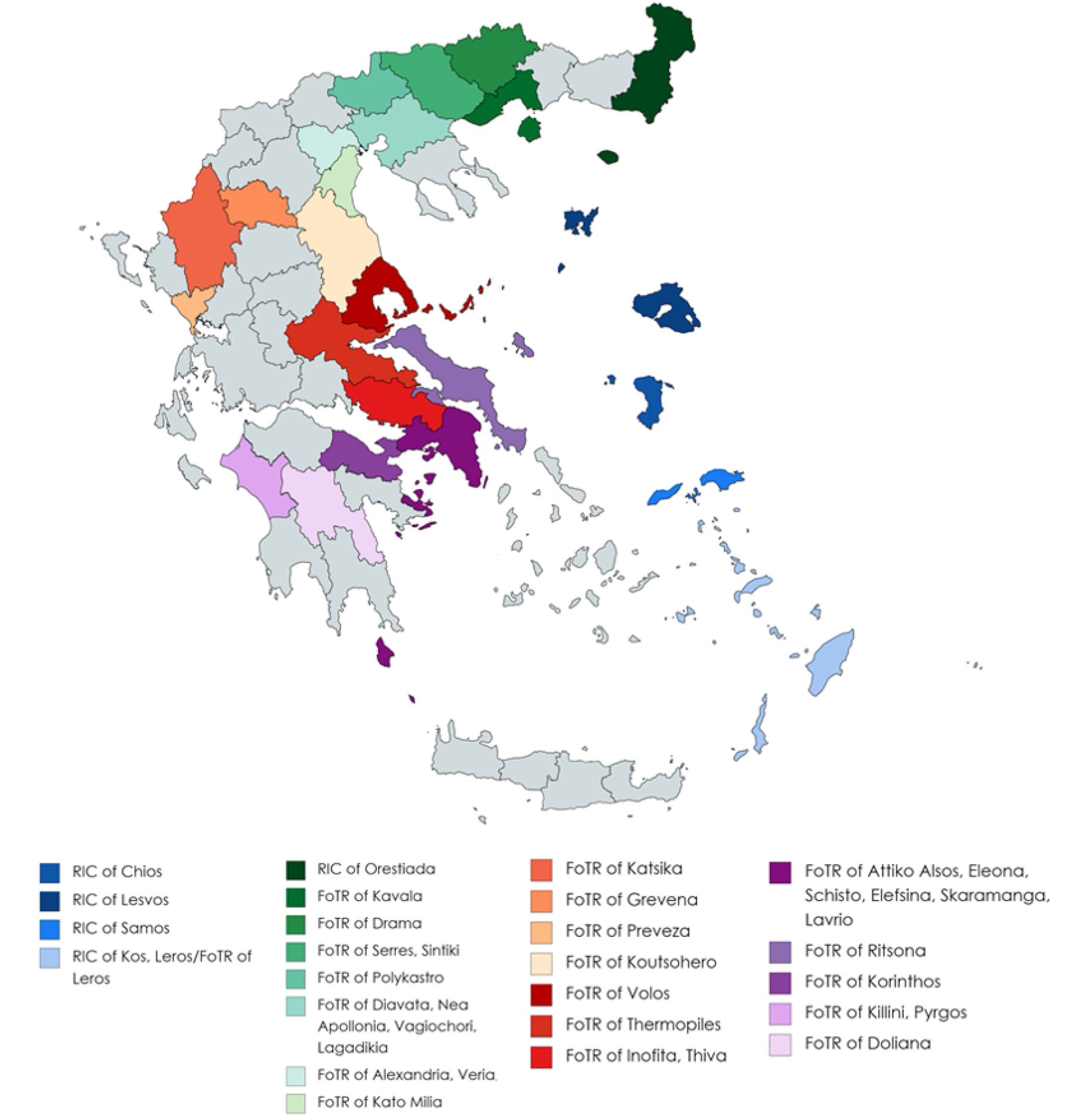
Geographic locations of Regional Units in Greece, as of November 2020. RIC= Reception and Identification Centers, FoTR= Facilities of Temporary Reception. Map created using MapChart (available from: https://mapchart.net/index.html)

### Scabies diagnosis

Diagnosis of scabies in PoC for refugees/asylum seekers of the NPHO is based on clinical skin examination, symptoms, and contact history. No laboratory confirmation is performed. This diagnosis is similar to the diagnosis of “Clinical Scabies,” in the consensus criteria developed by the International Alliance for the Control of Scabies in 2020 [11], although these criteria were not used specifically in this study.

### Data collection

Data was collected through the NPHO surveillance system in PoC for refugees/asylum seekers. PoC for refugees/asylum seekers include different health care facilities providing medical care within the refugees/asylum seekers centers and/or within hospitals and public primary care health facilities and serve as the first line of care for the people residing within the centers. NPHO systematically monitors 14 defined syndromes or health conditions, including respiratory infection with fever, gastroenteritis without blood in the stool, gastroenteritis with blood in the stool, rash with fever, clinical scabies, pulmonary tuberculosis, malaria with positive rapid diagnostic test, diphtheria, meningitis and/or encephalitis, hemorrhagic manifestations with fever and sepsis or septic shock.

Cumulative data is collected and sent daily from the participating PoC for refugees/asylum seekers. Data is collected by doctors, nurses and other health professionals working at the refugees/asylum seekers centers and Non-Governmental Organizations staffing primary care facilities. Participation rate (i.e., the number of centers sending data to NHPO), differs on a weekly basis. After analysis, detailed data is published in weekly reports online [12]. No individual clinical data is collected or published. The total population of refugees/asylum seekers at each center, and over time, is not reported.

### Treatment and management of scabies

Treatment instructions and medication supplies for scabies are under the authority of the Vector-mediated Diseases department of the NPHO. All treatment and infection control measures are implemented in collaboration with the NPHO, the Ministry of Migration and Asylum and other organizations responsible for refugee/migrant health care.

Treatment is administered to all suspected scabies cases and their close contacts. NPHO recommends application of benzyl benzoate lotion for adults and older children and precipitated sulfur vaseline for infants, young children and pregnant women. Dosages and duration of treatment differs per case according to the treating doctor’s assessment. Prophylactic mass administration of ivermectin or any other scabies treatment is not currently used systematically in the refugee/asylum seekers centers, but has been used to control outbreaks and/or clusters.

In addition, along with the administration of the medication for scabies, NPHO proposes a bundle of environmental measures. Use of clean clothes and bed sheets is recommended after, at least, the first two applications of the treatment. Clothes and bed sheet should be washed at high temperatures and thoroughly dried afterwards. When this is not feasible, clothes and bed sheets should be enclosed in a bag for seven days. Thorough cleaning of the living area with standard cleaning detergents is recommended but sterilization of the living environment or application of insecticides is not recommended.

### Statistical analysis

Data were analyzed by month for the period June 2016 to July 2020. The proportional morbidity for scabies was calculated as the proportion of cases of scabies compared to total consultations for all infective causes. The number of cases and proportional morbidity for four other syndromes (gastroenteritis without blood in the stool, respiratory infections, tuberculosis, and rash with fever) were also analyzed, to provide an overview of the health status of the population over time and a comparison for the trends of scabies cases. Descriptive analyses were performed using Microsoft Excel. Spearman correlation test was performed, using RStudio and R for Windows 4.0.2, to investigate scabies cases through time.

### Ethics

All data included in the study are publicly available through the official site of NPHO and do not include identifiable information. The study was approved by the Greek NPHO (KΠ 22069/2020-19/10/2020).

## Results

Data was collected through the system of epidemiological surveillance in PoC for refugees/asylum seekers of NPHO from June 20, 2016 to July 26, 2020, with the exception only of week 29 of 2017, for which no data were available (in total, 213 weeks of data were included). The number of refugees/asylum seekers centers sending data to the epidemiological surveillance system differed per week. The median weekly response rate was 95.2% (40 out of 42 PoC), with a minimum of 68% (week 52, 2018, 17 out of 25 PoC). There was a complete response (100%) for 53 weeks of the study.

In total, 1054807 consultations were documented for all causes. The median number of consultations, per month, was 19560 (IQR 17274 – 23549, Figure 2).

**Figure 2.**
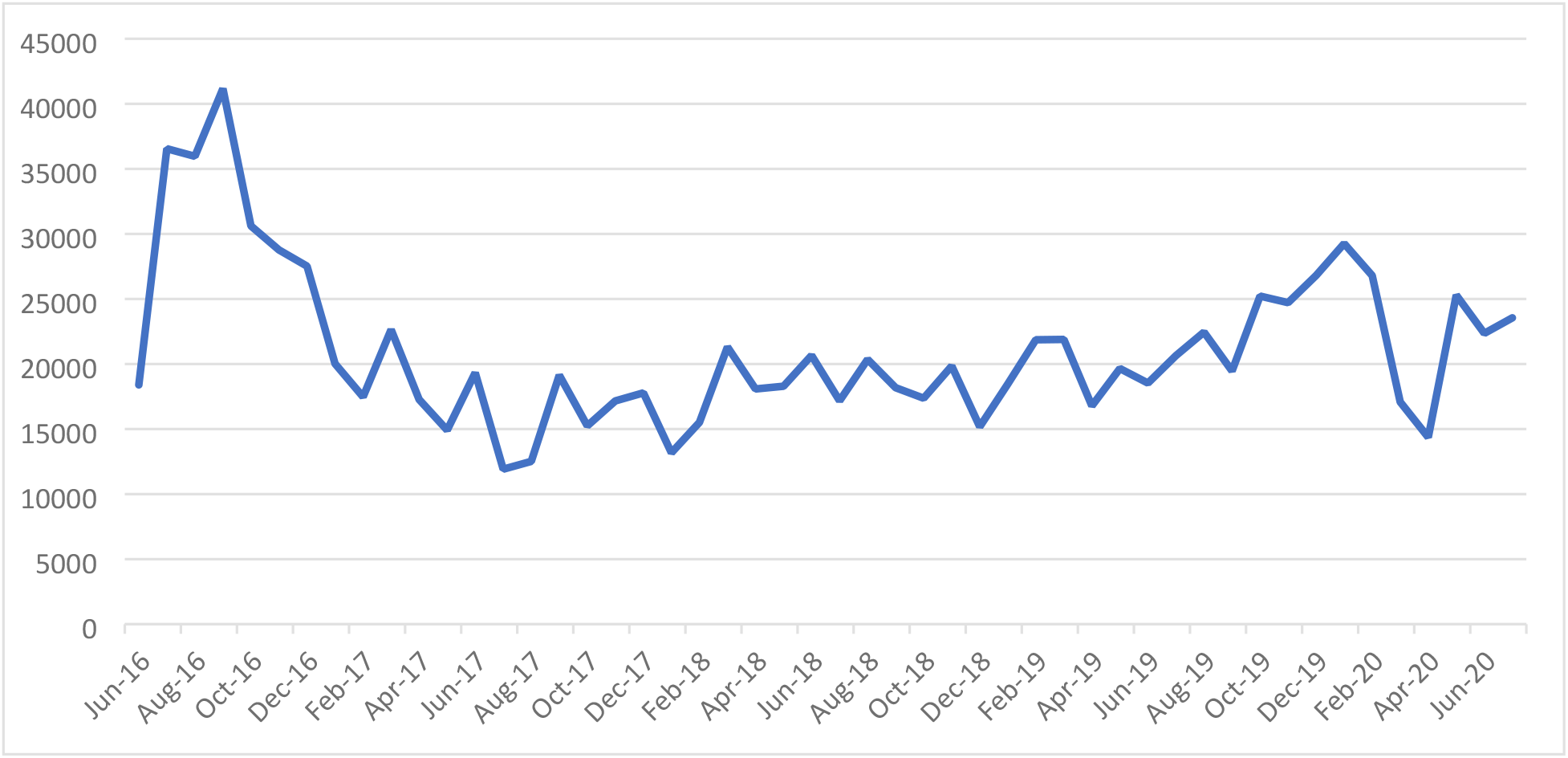
Total reported consultations at the centers hosting refugees/asylum seekers, for all causes, per month.

### Scabies epidemiology

A total of 13118 consultations with clinical scabies were reported over the study period (median 176, IQR 117 – 292, Figure 3). A maximum number of 1663 cases was observed in January 2020, related to a large outbreak.

**Figure 3.**
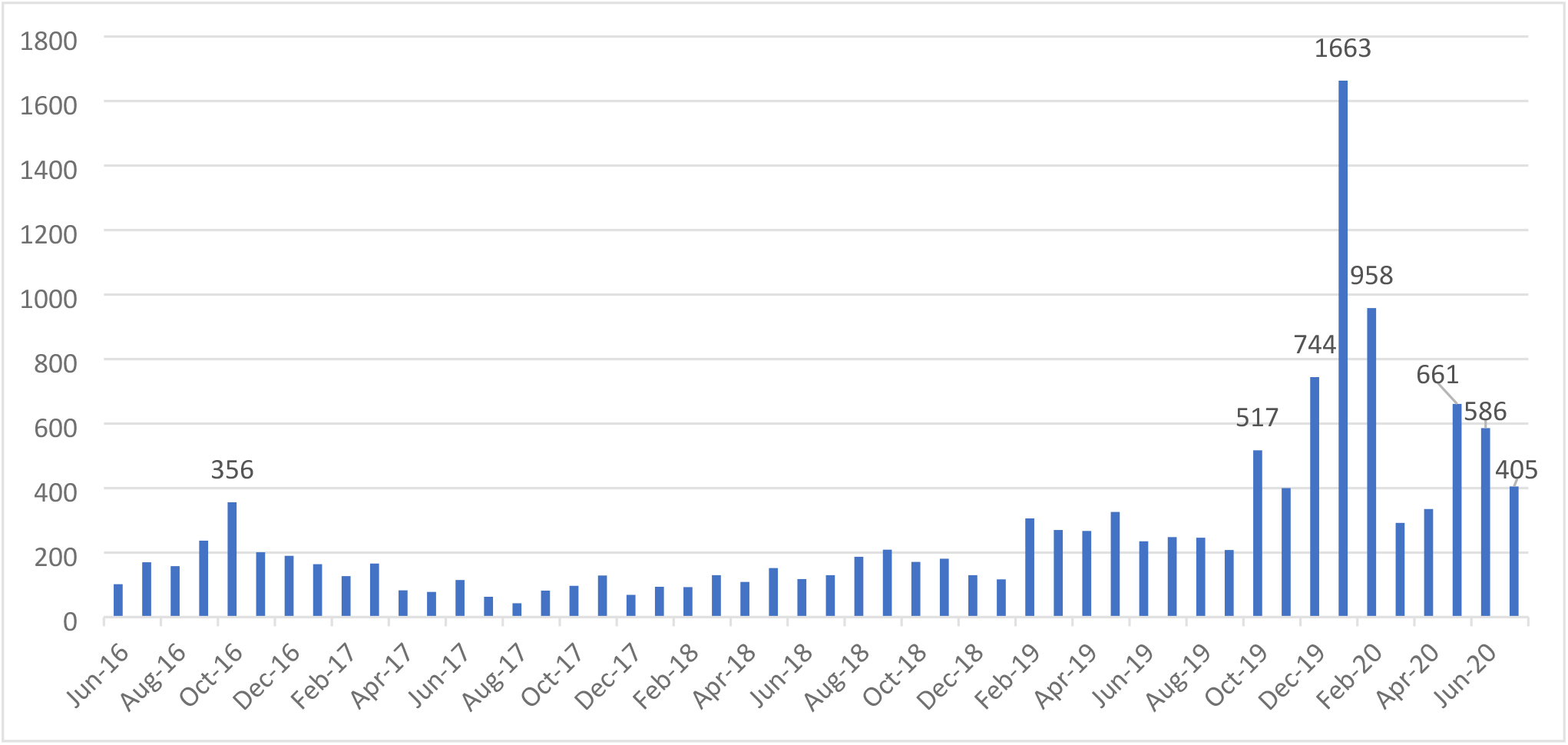
Number of consultations for clinical scabies reported by centers hosting refugees/asylum seekers, per month.

Although scabies cases fluctuated throughout the study period, the number of cases significantly increased from October 2019 until the end of the study period, with a peak of 1663 cases in January 2020. Spearman correlation test between reported scabies cases and time confirmed an increasing trend over the study period and a strong correlation between the two variables (R=0.67, Figure 4). The correlation remained strong (R=0.65) even after removing the January 2020 peak.

**Figure 4.**
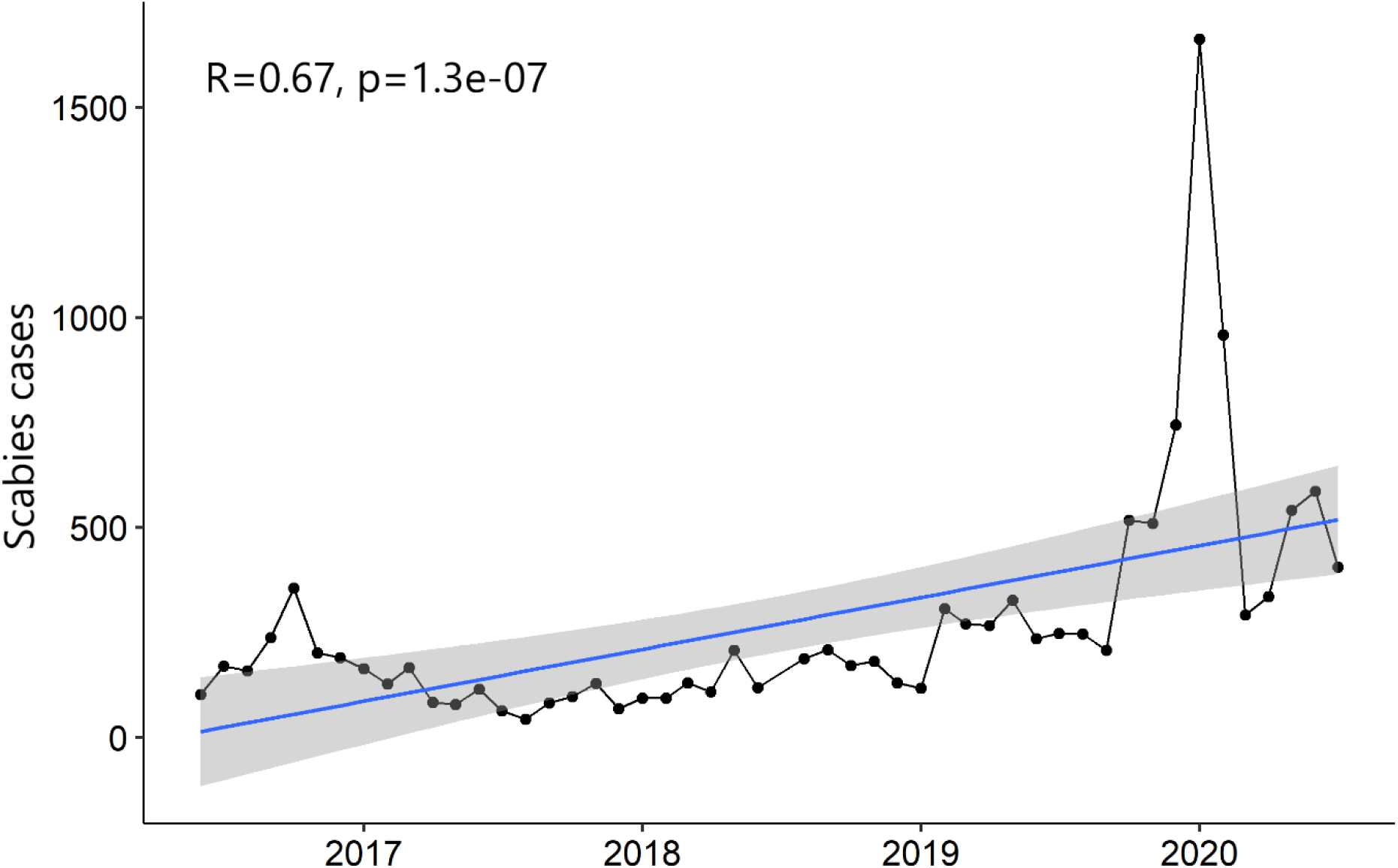
Trend of reported scabies cases during the time of the study period, including all data.

The proportion of consultations with scabies varied from 0.3% in August 2017 to 5.6 % in January 2020. The number of consultations for clinical scabies cases compared to other syndromes is shown in Figure 5. Respiratory infections, gastroenteritis without blood and scabies were the most frequent causes for consultations amongst the syndromes included in the NHPO epidemiological surveillance system. The observed peaks in consultations for these syndromes did not follow a specific seasonal pattern. However, consultations numbers peaked during the summer of 2016 and winter of 2020. The number of suspected tuberculosis cases remained relatively low throughout the study period.

**Figure 5.**
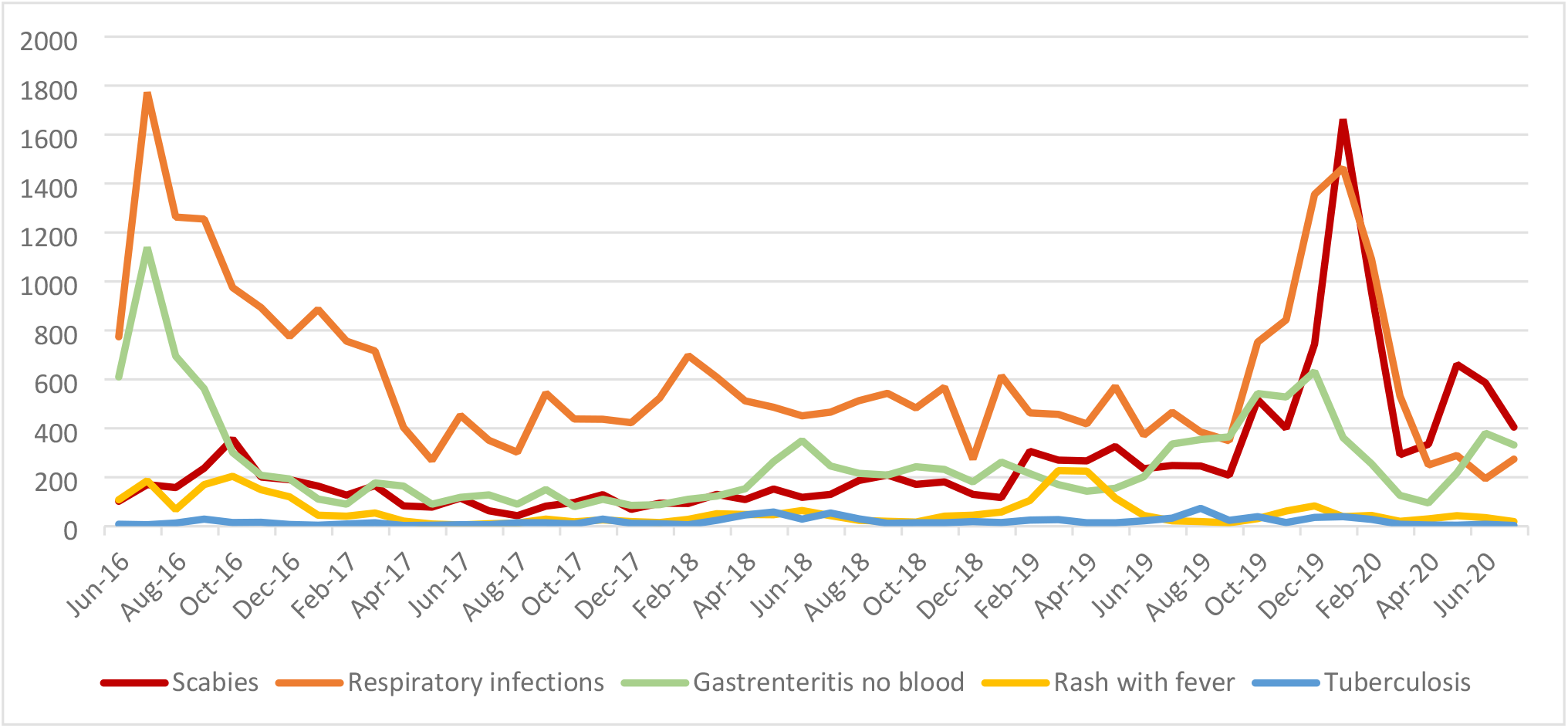
Number of consultations for clinical cases of scabies as compared to the four most common reported infectious diseases*, at the PoC, per month. *gastroenteritis without blood in the stool, respiratory infections, tuberculosis, rash with fever.

## Discussion

To our knowledge, this is the first study to investigate the epidemiology of scabies in the refugees/asylum seekers populations in Greece. Clinical scabies cases were documented for a four-year period from PoC for refugees/asylum seekers in Greece, with a very high average surveillance response rate.

Our study demonstrated that clinical scabies is a commonly reported infectious disease in this population. Scabies cases were reported from refugees/asylum seekers centers every week, and the number of consultations increased over the study period.

The increased number of clinical scabies cases during the first few weeks of 2020 is related to an outbreak of scabies observed in a PoC in a refugees/asylum seeker center. The specific center reported approximately 80% of the total scabies cases that month, mainly regarding newly arrived refugees, who manifested clinical symptoms before their arrival at the center [13].

Scabies is a major burden amongst vulnerable populations such as refugees/asylum seekers. Refugees/asylum seekers often live in crowded conditions throughout their transnational migrating route, increasing the transmission of scabies. In a 2020 study, regarding rescued people by non-governmental organization Open Arms in the Central Mediterranean, scabies was the most frequent infectious disease observed, accounting for 96% of total infectious diseases diagnoses [14]. Another study from Germany investigated infectious diseases among a cohort of unaccompanied refugee minors, and found scabies to be the most frequent infectious disease, affecting 14.2% of minors [15]. Furthermore, a systematic review that assessed the infectious disease profiles of Syrian and Eritrean migrants in Europe, found a high prevalence of scabies, up to 80% prevalence in the Eritrean migrants [16]. Patients with scabies, particularly children have to deal with the burden of the disturbing symptoms of the disease [17].

We report a very high burden of scabies among the refugees/asylum seekers population in Greece, with cases reported throughout the study period. Scabies was the third most frequent infectious disease, included in the systematically reported to NPHO syndromes. At specific time points, namely January 2020 and May-July 2020, scabies was the single most frequent reported infectious disease. Outbreaks of scabies cases coincided with peaks in other infectious diseases, with the most prominent example in January of 2020. A possible explanation could be an increase of the refugees/asylum seekers residing in the centers at the specific time points, the crowded conditions within the centers and/or seasonality of infectious diseases. No information is available on how many cases were complicated leading to secondary bacterial infections.

The rapid turnover of populations in the refugees/asylum seekers centers predisposes to reinfestations and outbreaks of scabies. A study from the Netherlands described mass drug administration of ivermectin/permethrin in Eritrean and Ethiopian refugees with a high prevalence of scabies. The number of reinfestations and the number of complicated forms of scabies reduced after mass drug administration [18]. Population-level mass drug interventions, particularly with oral ivermectin, can be more effective than individual case management, especially if scabies prevalence is over 5-10% [19]. Currently, in Greece, mass administration for prevention of scabies in refugees/asylum seekers is not implemented. Considering the increasing caseload of scabies and the crowded living conditions, mass drug administration, targeting newly-arrived refugees/asylum seekers may be an effective strategy. The current NPHO surveillance and response system could then act to detect and manage new cases and any emerging outbreaks in a timely and effective manner.

It is possible that infection control measures aiming to reduce the transmission of the SARS-CoV-2 virus may have influenced the number of consultations for infectious diseases under surveillance after March of 2020 [20,21]. However, respiratory and gastroenteritis infections cases in 2020 followed a similar pattern to the previous years, possibly reflecting the challenges with implementing social distancing in these refugee/asylum seekers centers.

Continuous, systematic data collection within the system of epidemiological surveillance in PoC for refugees/asylum seekers and subsequent publication by NPHO, provided valuable information on infectious diseases prevalence among the specific population allowing earlier interventions. Data were collected from a large number of PoC with wide geographical distribution, reflecting clear overview of scabies epidemiology in refugees/asylum seekers population in centers.

This study has several limitations. Data on complications of scabies, therapeutic adherence, clinical outcomes or reinfestation of scabies in Greece are not systematically documented and were therefore not available to include. Furthermore, demographic characteristics of the refugees/asylum seekers are not available, hence reported scabies cases could not be related to the country of origin. This study included only the scabies patients who visited the health care facilities. Therefore, the number of scabies included in our reports is likely to be an underestimation of the actual burden of scabies in the refugee centers.

### Conclusion

Scabies is one of the most frequent reported infectious diseases among the refugees/asylum seekers population in Greece. The number of consultations due to scabies increased over the period 2016 to 2020. National systematic reporting of scabies by health personnel working in PoC can enable early interventions in order to reduce the burden of scabies and its complications. As the incidence increases and living conditions complicate optimal treatment of individual patients, mass drug administration may prove to be a necessary intervention to reduce the burden of scabies in refugees/asylum seekers population.

## Data Availability

All data used for the analysis/findings of this manuscript is publicly available through the weekly reports of the official site of Greek National Public Health Organisation. Available from: https://eody.gov.gr/en/epidemiological-statistical-data/system-of-epidemiological-surveillance-in-points-of-care-for-refugees-migrants/

## Acknowledgements

We would like to acknowledge all medical staff responsible for refugees/asylum seekers health care at the Regional Units in Greece that contributed to the documentation and report of the data at the NPHO. Furthermore, we would like to thank the corresponding departments of NHPO that gave us access to relevant information.

